# Systolic Blood Pressure Time in Target Range and Cerebral Small Vessel Disease

**DOI:** 10.64898/2025.12.24.25342993

**Authors:** Guanming Ji, Mengmeng Bai, Huijing Shi, Ling Yang, Xianyu Zhu, Rui Li, Jing Li, Han Lv, Pengfei Zhao, Shuohua Chen, Yuntao Wu, Ying Hui, Shouling Wu, Zhenchang Wang, Xiaoshuai Li

## Abstract

**BACKGROUND:** Whether systolic blood pressure (SBP) time in target range (TTR) associates with cerebral small vessel disease (CSVD) in hypertensive patients remains unclear.

**METHODS:** Between December 2020 and August 2023, participants were prospectively included from the Kailuan study. TTR was calculated using linear interpolation with the SBP target range between 120 and 140 mm Hg from 2006 to 2020. Beginning in 2020, brain MRI was conducted to identify CSVD burden and imaging markers, namely, lacunes (LAs), cerebral microbleeds (CMBs), enlarged perivascular spaces (EPVSs), and white matter hyperintensities (WMHs). Logistic regression models were used to assess the association between SBP-TTR and imaging markers of CSVD.

**RESULTS:** A total of 797 patients with hypertension were included from the Kailuan Study. Compared with the lowest tertile, the highest tertile of SBP-TTR was associated with a reduced risk of EPVS (OR = 0.67; 95% CI: 0.47–0.96; P = 0.029) and WMHs (OR = 0.66; 95% CI: 0.44–0.97; P = 0.033). This association was independent of SBP variability. No significant associations were found between SBP-TTR and LAs (OR = 0.69; 95% CI: 0.43–1.12; P = 0.133) or CMBs (OR = 0.97; 95% CI: 0.66–1.43; P = 0.869). Subgroup analysis suggested that the protective effect of a higher SBP-TTR against EPVSs was more pronounced in men, participants aged ≥65 years, those with a follow-up duration of ≥5 years, and nondiabetic patients.

**CONCLUSIONS:** A higher SBP-TTR is significantly associated with a reduced risk of EPVSs and WMHs, independent of SBP variability.

## Introduction

As the global population ages, the prevalence of CSVD continues to increase, exceeding 70% among middle-aged and older adults^1,2^. CSVD is not only strongly associated with clinical manifestations, such as cognitive impairment, urinary and defecatory dysfunction, and gait abnormalities, but also substantially increases the risk of stroke, vascular dementia, and Alzheimer’s disease, thereby establishing it as a significant global health challenge^3,4^.

Hypertension represents the most important modifiable risk factor for CSVD^5^. Previous studies have indicated that each 1 standard deviation increase in systolic blood pressure (SBP) is associated with an approximately 25% increased risk of overall CSVD burden^6^. Clinical guidelines emphasize blood pressure control as the most effective strategy for preventing the onset and progression of age-related CSVD. However, blood pressure exhibits substantial temporal variability^7^. Relying solely on single or clinic-based measurements is insufficient for accurately assessing long-term blood pressure control. Recent evidence has suggested that blood pressure variability (BPV), particularly SBPV, is independently associated with CSVD burden ^8,9^. Hence, identifying a composite indicator that reflects both blood pressure level and variability is critical for evaluating the quality of blood pressure management and assessing CSVD risk.

In recent years, the time in target range (TTR) has been proposed as a novel metric for quantifying long-term blood pressure control^10,11^. TTR is defined as the proportion of time during which blood pressure remains within a predefined target range over the total follow-up period^12^. This metric integrates information regarding both average blood pressure levels and variability, as well as fluctuations both within and outside the target range^13^. Recent studies have demonstrated that the SBP-TTR is significantly associated with cardiovascular and cerebrovascular diseases and cognitive impairment^14,15^. However, the association between the SBP-TTR and CSVD burden in hypertensive patients remains unclear. On the basis of this background, the present study utilized multimodal neuroimaging data from the Kailuan Study to systematically investigate the association between SBP-TTR and CSVD burden. The aim of this investigation is to establish a more comprehensive paradigm for evaluating blood pressure management efficacy and to furnish novel empirical evidence informing the prevention and clinical management of CSVD.

## 1 Materials and Methods

### 1.1 Study population

The Kailuan Study is a prospective dynamic cohort study based on a large community in Tangshan, China. From July 2006 to October 2007, a total of 101,510 adult participants (≥18 years) were enrolled; baseline questionnaires, clinical examinations, and laboratory tests were completed, and biennial follow-ups were conducted thereafter. The study protocol and implementation procedures have previously been described in detail. The seventh follow-up began in December 2020, during which time a subset of participants concurrently underwent a brain magnetic resonance imaging (MRI) assessment.

From December 2020 to August 2023, a total of 1,900 participants participated in this follow-up. The inclusion criteria were as follows: (1) participants who were diagnosed with hypertension at the 2006 baseline examination, who had a prior history of hypertension, or who developed new-onset hypertension during follow-up between 2008 and 2018; and (2) participants who completed the brain MRI examination in 2020. Exclusion criteria: Participants who had a history of stroke or tumor, who had incomplete imaging data, or who had missing SBP data were excluded. Finally, 797 participants were included in this study. The study protocol was approved by the Ethics Committee of Kailuan General Hospital. Written informed consent was obtained from all participants.

### 1.2 Blood Pressure Measurement and Calculation of TTR

Blood pressure measurements were performed by trained healthcare personnel using a standardized procedure. After the participants had rested quietly for 5 min in a seated position, their blood pressure was measured in their left upper arm. Three consecutive measurements were taken at 1- to 2-min intervals, and the average of these three readings was used for statistical analysis. In accordance with current clinical guidelines, the target range for SBP was defined as 120–140 mmHg in this study.

The time span from the year of hypertension diagnosis for each participant until the year of the MRI examination in 2020 was defined as the observation window. This window was divided into seven consecutive subintervals based on the biennial examination years (2006–2008; 2008–2010; 2010–2012; 2012–2014; 2014–2016; 2016–2018; 2018–2020). For individual participants, a specific combination of these subintervals, starting from their year of hypertension diagnosis, constituted their actual observation window. Within each subinterval, linear interpolation was applied to the SBP values measured at adjacent examinations to approximate the blood pressure trajectory over time. The SBP status throughout the subinterval was subsequently assessed: if the SBP consistently resided within the target range for the entire subinterval, the full subinterval duration was classified as time in range; conversely, if the SBP was within the target range for only a portion of the subinterval, the precise duration was computed based on the interpolation. The cumulative time within the target range across all subintervals was then aggregated, divided by the total observation window duration, and expressed as a percentage value. This metric was defined as the SBP time in target range (SBP-TTR, %).

### 1.3 MRI Data Acquisition

Conventional MRI sequences included T1-weighted imaging (T1WI), T2-weighted imaging (T2WI), T2-fluid attenuated inversion recovery (FLAIR), susceptibility-weighted imaging (SWI), and axial diffusion-weighted imaging (DWI).

The scanning parameters were as follows: T1WI: TE/TR = 7.8/500 ms; FA = 90°; slice thickness = 5 mm; FOV = 256 mm × 256 mm. T2WI: TE/TR = 7/4000 ms; FA = 150°; slice thickness = 5 mm; FOV = 256 mm × 256 mm. FLAIR: TE/TR = 1147/5000 ms; FA = 90°; slice thickness = 1 mm; FOV = 256 mm × 256 mm. SWI: TE/TR = 40/26 ms; slice thickness = 2 mm; FOV = 240 mm × 240 mm. DWI: TE/TR = 5110/77.2 ms; FA = 90°; slice thickness = 5 mm; FOV = 240 mm × 240 mm.

### 1.4 Cerebral Small Vessel Disease Assessment

Lacunes (LAs) were operationally defined on MRI as round or ovoid cerebrospinal fluid (CSF)-like signal cavities located within subcortical regions. Their characteristic radiological features included hypointensity on T1WI, hyperintensity on T2WI, and a distinctive presentation on FLAIR sequences featuring a central hypointense signal surrounded by a hyperintense rim. The diameters of the lacunes ranged from 3 to 15 mm. Cerebral microbleeds (CMBs) were defined as small, well-demarcated, homogeneous, round foci exhibiting signal voids on SWI, attributable to the paramagnetic effect of hemosiderin deposits. Conversely, CMBs were not discernible on FLAIR, T1WI, or T2WI sequences. Enlarged perivascular spaces (EPVSs) were identified as CSF-like signal spaces tracing the course of penetrating cerebral vessels. These spaces manifested as hypointensity on T1WI and FLAIR sequences and hyperintensity on T2WI. EPVSs typically have diameters <3 mm and are predominantly located within the basal ganglia, subcortical white matter, and brainstem. White matter hyperintensity (WMH) refers to regions of abnormal signal intensity within the white matter that appear hyperintense on both T2WI and FLAIR sequences but demonstrate isointensity or hypointensity on T1WI.

The severity of WMHs was assessed using the Fazekas scale, which combines scores for periventricular WMHs (PVWMHs) and deep WMHs (DWMHs), yielding a total score ranging from 0 to 6^16^. The severity of EPVS was graded using a 4-point scale based on the slice with the greatest involvement in the basal ganglia. The total radiological burden score of CSVD was defined as follows. One point was assigned for the presence of any one of the following features: the presence of ≥1 lacune; a Fazekas score of ≥2 for DWMHs and/or a score of 3 for PVWMH; the presence of ≥1 deep or infratentorial microbleed; and moderate to severe EPVSs (>10) in the basal ganglia^17^.(Figure 1)

**Figure 1:**
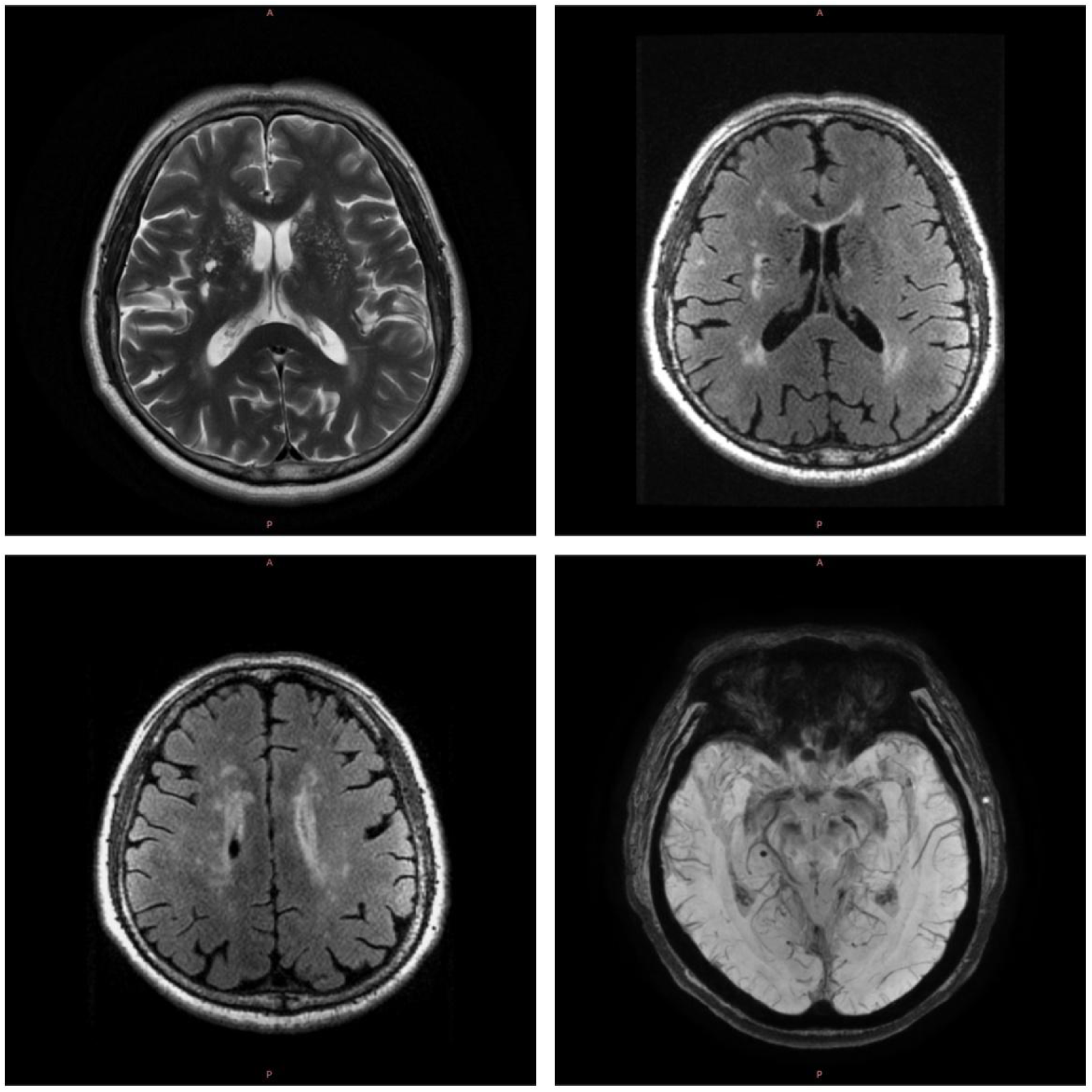

The total CSVD burden score ranged from 0 to 4. On the basis of previous research^18^, the CSVD score was dichotomized in this study: a score of 0 was defined as “no CSVD burden,” and a score of ≥1 was defined as “presence of CSVD burden.”

### 1.5 Covariates

Information on age, sex, alcohol consumption history, smoking history, physical activity, medical history (diabetes, hypertension, and dyslipidemia), and medication use (antihypertensive, hypoglycemic, and lipid-lowering drugs) was collected via questionnaires. Body mass index (BMI) was calculated from height and weight measurements. Hypertension was defined as a blood pressure ≥140/90 mmHg or current use of antihypertensive medication. Diabetes mellitus was defined as a fasting blood glucose level ≥7.0 mmol/L or the use of hypoglycemic medication. Dyslipidemia was defined as a low-density lipoprotein cholesterol (LDL-C) concentration ≥4.1 mmol/L, a high-density lipoprotein cholesterol (HDL-C) concentration ≤1.0 mmol/L, a triglyceride (TG) concentration ≥2.3 mmol/L, or a total cholesterol (TC) concentration ≥6.2 mmol/L.

### 1.6 Statistical Analysis

All the statistical analyses were performed using SAS software, version 9.4 (SAS Institute Inc., Cary, NC, USA). A two-sided P value < 0.05 was considered to indicate statistical significance. The baseline characteristics of the participants were calculated according to SBP-TTR tertiles.

Continuous variables were assessed for normality. Normally distributed variables are expressed as the means ± standard deviations and were compared using one-way analysis of variance (ANOVA). Nonnormally distributed variables are expressed as medians (interquartile ranges) and were compared using the Kruskal‒Wallis test. Categorical variables are expressed as frequencies (%) and were compared using the χ² test. Missing values for covariates were handled using multiple imputation. Logistic regression models were used to evaluate the associations between SBP-TTR and total CSVD burden, LAs, CMBs, EPVSs, and WMH, generating odds ratios (ORs) with corresponding 95% confidence intervals (CIs). Two adjusted models were constructed: Model 1: Adjusted for sex and age. Model 2: Adjusted for Model 1 covariates plus smoking history, alcohol consumption history, physical activity level, BMI, baseline SBP, fasting blood glucose level, LDL-C level, antihypertensive medication use, glucose-lowering medication use, and lipid-lowering medication use.

Additionally, stratified analyses were conducted according to follow-up window (≥5 years vs. <5 years), age (<65 years vs. ≥65 years), sex, and diabetes status to explore potential differences among the subgroups.

To evaluate the robustness of the results, blood pressure variability indices, namely, the standard deviation of SBP (SBP-SD) and the coefficient of variation of SBP (SBP-CV), were included in the fully adjusted model (Model 2) to examine the impact of blood pressure fluctuations on the association.

## 2. Results

### 2.1 Baseline Characteristics of the Study Participants

A total of 797 hypertensive participants meeting the inclusion criteria were ultimately included in this study. The mean age at the time of MRI examination was 56.71 ± 11.12 years (range: 30.5–81.8 years), and 447 (59.8%) were men. The mean follow-up window for all participants was 10.09 years. Table 1 presents the demographic characteristics, clinical indicators, and radiological characteristics of all participants at the end of follow-up, stratified by SBP-TTR tertiles. No statistically significant differences were observed in the aforementioned characteristics among the different SBP-TTR groups.

**Table 1.** Baseline characteristics of participants based on systolic blood TTR.

|  | Total | SBP-TTR1<br>< 17% | SBP-TTR2<br>17%- < 57% | SBP-TTR3<br>≥57% | *p*-Value |
| --- | --- | --- | --- | --- | --- |
| Participants, n | 797 | 265 | 266 | 266 |  |
| Age, year | 56.71±11.12 | 57.13±10.92 | 56.61±11.34 | 56.4±11.12 | 0.736<sup>a</sup> |
| Male, n (%) | 477 (59.8) | 159 (60.0) | 147 (55.3) | 171 (64.3) | 0.105<sup>c</sup> |
| Smoking history, n (%) | 286 (35.9) | 99 (37.4) | 86 (32.3) | 101 (38.0) | 0.331<sup>c</sup> |
| Drinking history, n (%) | 357 (44.8) | 123 (46.4) | 104 (39.1) | 130 (48.9) | 0.062<sup>c</sup> |
| Physical exercise, n (%) | 512 (64.2) | 170 (64.2) | 168 (63.2) | 174 (65.4) | 0.862<sup>c</sup> |
| BMI, kg/m<sup>2</sup> | 25.30±3.56 | 25.42±3.53 | 24.98±3.3 | 25.51±3.83 | 0.186<sup>a</sup> |
| SBP, mm/Hg | 134.67±19.76 | 135.91±19.76 | 132.83±19.68 | 135.27±19.78 | 0.166<sup>a</sup> |
| FBG, mmol/L | 5.0 (4.69,5.69) | 5.06 (4.70,5.65) | 5.03 (4.66,5.64) | 5.14 (4.73,5.95) | 0.068<sup>b</sup> |
| LDL-C, mmol/L | 3.13±0.77 | 3.11±0.76 | 3.1±0.77 | 3.18±0.79 | 0.434<sup>a</sup> |
| Diabetes, n(%) | 152 (19.1) | 51 (19.3) | 50 (18.8) | 51 (19.2) | 0.990<sup>c</sup> |
| Hyperlipidemia, n (%) | 451 (56.6) | 159 (60.0) | 139 (52.3) | 153 (57.5) | 0.184<sup>c</sup> |
| Antihypertensive treatment, n (%) | 311 (39.0) | 120 (45.3) | 97 (36.5) | 94 (35.3) | 0.037<sup>c</sup> |
| Hypoglycemic treatment, n (%) | 83 (10.4) | 20 (7.6) | 26 (9.8) | 37 (13.9) | 0.051<sup>c</sup> |
| Lipid-lowering treatment, n (%) | 124 (15.6) | 52 (19.6) | 37 (13.9) | 35 (13.2) | 0.080<sup>c</sup> |
| TMBC, n (%) | 556 (69.8) | 194 (73.2) | 186 (69.9) | 176 (66.2) | 0.210<sup>c</sup> |
| LA, n (%) | 140 (17.6) | 50 (18.9) | 53 (19.9) | 37 (13.9) | 0.151<sup>c</sup> |
| CMB, n (%) | 229 (28.7) | 75 (28.3) | 77 (29.0) | 77 (29.0) | 0.982<sup>c</sup> |
| EPVS, n (%) | 477 (59.8) | 172 (64.9) | 158 (59.4) | 147 (55.3) | 0.075<sup>c</sup> |
| WMH, n (%) | 226 (28.4) | 87 (32.8) | 73 (27.4) | 66 (24.8) | 0.113<sup>c</sup> |

Values are mean SD , median (interquartile range) or n (%). BMI,body mass index; SBP, Systolic Blood Pressure; FBG, Fasting Blood Glucose; LDL-C, Low-Density Lipoprotein Cholesterol; TMBC, Total burden of cerebral small vessel disease; LA, Lacunes; CMB, Cerebral Microbleeds; EPVS, Enlarged Perivascular Spaces; WMH, White Matter Hyperintensities.
a one-way analysis of variance
b Wilcoxon Rank Sum Test
c chi-square test

### 2.2 Associations between SBP-TTR and CSVD

The associations between SBP-TTR and CSVD are shown in Table 2. Compared with the lowest tertile, the highest SBP-TTR tertile was associated with a reduced risk of EPVSs (OR = 0.67; 95% CI: 0.47–0.96; P = 0.029) and WMHs (OR = 0.66; 95% CI: 0.44–0.97; P = 0.033). No significant associations were observed with total CSVD burden (OR = 0.71; 95% CI: 0.49–1.04; P = 0.083), LAs (OR = 0.69; 95% CI: 0.43–1.12; P = 0.133) or CMBs (OR = 0.97; 95% CI: 0.66–1.43; P = 0.869).

**Table 2.**
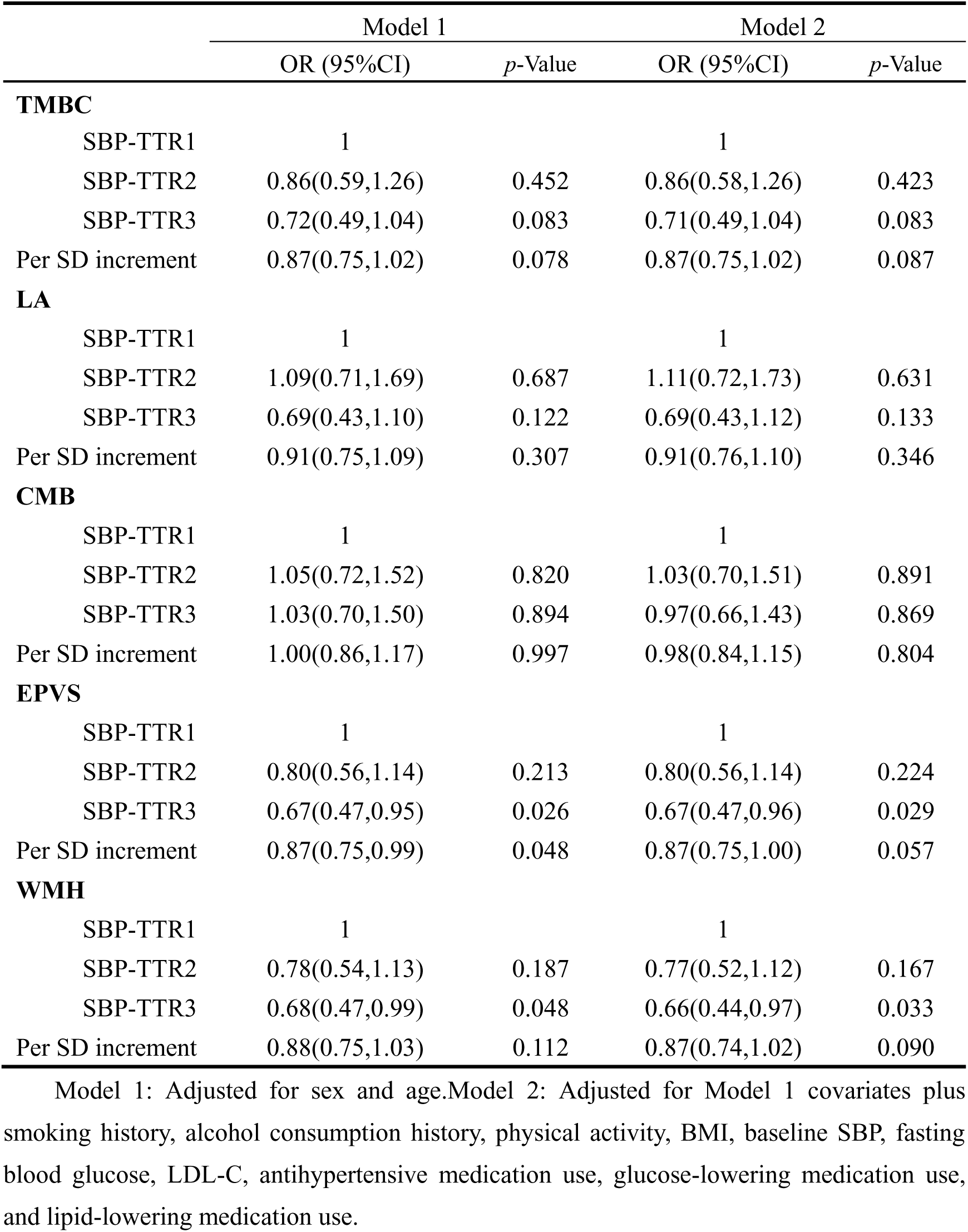
Association of TTR of SBP with CSVD burden.

|  | Model 1 |  | Model 2 |  |
| --- | --- | --- | --- | --- |
|  | OR (95%CI) | *p*-Value | OR (95%CI) | *p*-Value |
| **TMBC** |  |  |  |  |
| SBP-TTR1 | 1 |  | 1 |  |
| SBP-TTR2 | 0.86(0.59,1.26) | 0.452 | 0.86(0.58,1.26) | 0.423 |
| SBP-TTR3 | 0.72(0.49,1.04) | 0.083 | 0.71(0.49,1.04) | 0.083 |
| Per SD increment | 0.87(0.75,1.02) | 0.078 | 0.87(0.75,1.02) | 0.087 |
| **LA** |  |  |  |  |
| SBP-TTR1 | 1 |  | 1 |  |
| SBP-TTR2 | 1.09(0.71,1.69) | 0.687 | 1.11(0.72,1.73) | 0.631 |
| SBP-TTR3 | 0.69(0.43,1.10) | 0.122 | 0.69(0.43,1.12) | 0.133 |
| Per SD increment | 0.91(0.75,1.09) | 0.307 | 0.91(0.76,1.10) | 0.346 |
| **CMB** |  |  |  |  |
| SBP-TTR1 | 1 |  | 1 |  |
| SBP-TTR2 | 1.05(0.72,1.52) | 0.820 | 1.03(0.70,1.51) | 0.891 |
| SBP-TTR3 | 1.03(0.70,1.50) | 0.894 | 0.97(0.66,1.43) | 0.869 |
| Per SD increment | 1.00(0.86,1.17) | 0.997 | 0.98(0.84,1.15) | 0.804 |
| **EPVS** |  |  |  |  |
| SBP-TTR1 | 1 |  | 1 |  |
| SBP-TTR2 | 0.80(0.56,1.14) | 0.213 | 0.80(0.56,1.14) | 0.224 |
| SBP-TTR3 | 0.67(0.47,0.95) | 0.026 | 0.67(0.47,0.96) | 0.029 |
| Per SD increment | 0.87(0.75,0.99) | 0.048 | 0.87(0.75,1.00) | 0.057 |
| **WMH** |  |  |  |  |
| SBP-TTR1 | 1 |  | 1 |  |
| SBP-TTR2 | 0.78(0.54,1.13) | 0.187 | 0.77(0.52,1.12) | 0.167 |
| SBP-TTR3 | 0.68(0.47,0.99) | 0.048 | 0.66(0.44,0.97) | 0.033 |
| Per SD increment | 0.88(0.75,1.03) | 0.112 | 0.87(0.74,1.02) | 0.090 |

Model 1: Adjusted for sex and age. Model 2: Adjusted for Model 1 covariates plus smoking history, alcohol consumption history, physical activity, BMI, baseline SBP, fasting blood glucose, LDL-C, antihypertensive medication use, glucose-lowering medication use, and lipid-lowering medication use.

### 2.3 Subgroup Analyses

The results of the subgroup analyses indicated that among participants whose observation window exceeded 5 years, a higher SBP-TTR was associated with a reduced risk of EPVSs. For each 1- standard deviation (SD) increase in the SBP-TTR, the risk of EPVSs decreased by 15% (adjusted OR = 0.85; 95% CI: 0.72–0.99; P = 0.044). No statistically significant associations were determined between SBP-TTR and WMHs, LAs, or CMBs (Figure 2). In participants with an observation window of less than 5 years, SBP-TTR was not significantly associated with EPVSs, WMHs, LAs, or CMBs (Figure 2). (Supplemental Material)

**Figure 2:**
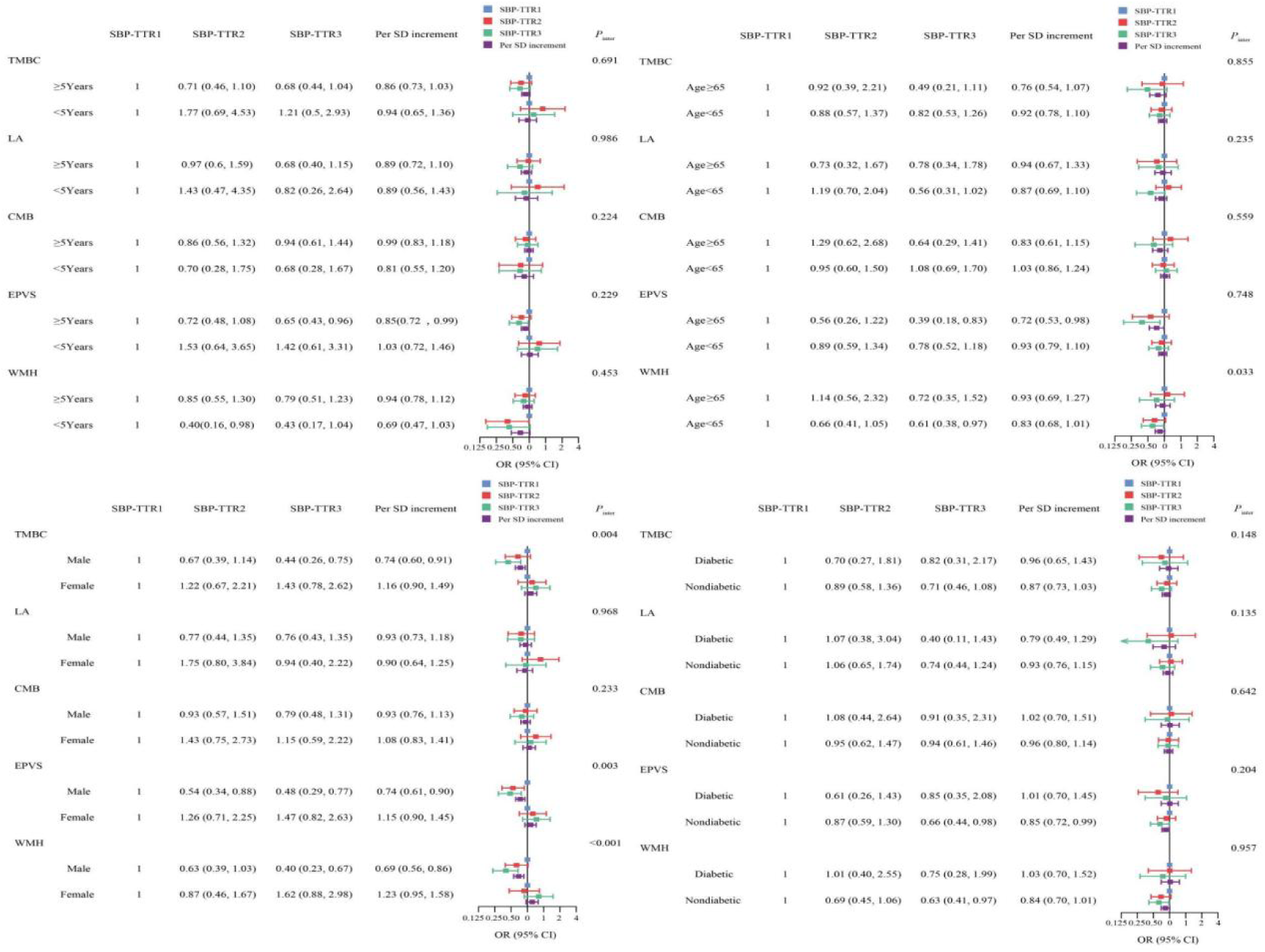

In the age-stratified analysis, a high SBP-TTR was associated with a reduced risk of EPVSs in participants older than 65 years (OR = 0.39; 95% CI: 0.18–0.83; P = 0.015) (Figure 2). Furthermore, subgroup analyses revealed that among men, SBP-TTR was negatively associated with both EPVSs (OR = 0.48, 95% CI: 0.29–0.77; P = 0.003) and WMHs (OR = 0.40, 95% CI: 0.23–0.67; P < 0.001) (Figure 2). Additionally, a significant negative association was observed between SBP-TTR and EPVSs in hypertensive participants without diabetes (OR = 0.66, 95% CI: 0.44–0.98; P = 0.038) (Figure 2). (Supplemental Material)

### 2.4 Sensitivity Analyses

After incorporating SBP variability metrics into the logistic regression models, the effects of SBP-TTR on the risks of EPVSs and WMHs were independent of SBP variability. After adjustment for SBP-SD, an increase in SBP-TTR was significantly associated with a reduced risk of EPVSs (OR = 0.67, 95% CI: 0.47–0.96, P = 0.029), and was also associated with a reduced risk of WMHs (OR = 0.65, 95% CI: 0.44–0.96, P = 0.031). After adjustment for the SBP-CV, an increase in the SBP-TTR remained significantly associated with a reduced risk of EPVSs (OR = 0.67, 95% CI: 0.47–0.96, P = 0.028) and was associated with a reduced risk of WMHs (OR = 0.65, 95% CI: 0.44–0.96, P = 0.031) (Table 3).

**Table 3.** Association of TTR of SBP with CSVD burden after adjustment for SBP variability metrics.

|  | SBP-std |  | SBP-cv |  |
| --- | --- | --- | --- | --- |
|  | OR (95%CI) | *p*-Value | OR (95%CI) | *p*-Value |
| **TMBC** |  |  |  |  |
| SBP-TTR1 | 1 |  | 1 |  |
| SBP-TTR2 | 0.86 (0.58, 1.26) | 0.431 | 0.86 (0.58, 1.26) | 0.432 |
| SBP-TTR3 | 0.72 (0.49, 1.05) | 0.085 | 0.71 (0.49, 1.05) | 0.083 |
| Per SD increment | 0.88 (0.75, 1.02) | 0.091 | 0.88 (0.75, 1.02) | 0.089 |
| **LA** |  |  |  |  |
| SBP-TTR1 | 1 |  | 1 |  |
| SBP-TTR2 | 1.11 (0.72, 1.73) | 0.630 | 1.10 (0.71, 1.71) | 0.663 |
| SBP-TTR3 | 0.69 (0.43, 1.11) | 0.126 | 0.69 (0.43, 1.11) | 0.127 |
| Per SD increment | 0.91 (0.75, 1.10) | 0.323 | 0.91 (0.75, 1.10) | 0.330 |
| **CMB** |  |  |  |  |
| SBP-TTR1 | 1 |  | 1 |  |
| SBP-TTR2 | 1.03 (0.70, 1.51) | 0.872 | 1.03 (0.70, 1.51) | 0.876 |
| SBP-TTR3 | 0.97 (0.66, 1.42) | 0.863 | 0.97 (0.66, 1.42) | 0.864 |
| Per SD increment | 0.98 (0.84, 1.15) | 0.809 | 0.98 (0.84, 1.15) | 0.810 |
| **EPVS** |  |  |  |  |
| SBP-TTR1 | 1 |  | 1 |  |
| SBP-TTR2 | 0.80 (0.56, 1.15) | 0.223 | 0.80 (0.56, 1.14) | 0.222 |
| SBP-TTR3 | 0.67 (0.47, 0.96) | 0.029 | 0.67 (0.47, 0.96) | 0.028 |
| Per SD increment | 0.87 (0.75, 1.00) | 0.057 | 0.87 (0.75, 1.00) | 0.056 |
| **WMH** |  |  |  |  |
| SBP-TTR1 | 1 |  | 1 |  |
| SBP-TTR2 | 0.76 (0.52, 1.11) | 0.158 | 0.76 (0.52, 1.11) | 0.153 |
| SBP-TTR3 | 0.65 (0.44, 0.96) | 0.031 | 0.65 (0.44, 0.96) | 0.031 |
| Per SD increment | 0.87 (0.74, 1.02) | 0.083 | 0.87 (0.74, 1.02) | 0.084 |

SBP-std, Standard Deviation of Systolic Blood Pressure; SBP-CV, Coefficient of Variation of Systolic Blood Pressure.

## 3. Discussion

In this study, on the basis of Kailuan community participants, the association between SBP-TTR and the risk of CSVD was investigated. The findings demonstrate that a higher SBP-TTR is significantly associated with a reduced risk of EPVSs and WMHs, and this association is independent of SBP variability. Subgroup analyses further revealed that the protective effect of a high SBP-TTR against EPVSs was more pronounced among male participants aged >65 years. Extending the time window during which the SBP is maintained within the target range may more effectively reduce the risk of EPVSs. These findings provide new intervention strategies and potential targets for the prevention and control of CSVD risk.

This study revealed that a high SBP-TTR significantly reduces the risk of enlarged perivascular spaces (EPVSs), independent of SBP variability. Perivascular spaces (PVSs), the fluid-filled spaces surrounding small penetrating arteries/arterioles and veins/venules as they traverse the subarachnoid space and brain parenchyma, serve as critical conduits for metabolic clearance via the glymphatic system^19^. Dysfunction of the glymphatic system, characterized by impaired fluid dynamics, can lead to obstructed fluid flow within the PVSs, resulting in fluid accumulation and ultimately PVS enlargement ^20,21^.

Studies have confirmed that enlarged PVSs are closely associated with neurodegenerative diseases, cognitive impairments, dementia, and sleep disorders^19,22^. A community-based investigation conducted by Shi et al. demonstrated that cumulative exposure to high SBP, diastolic blood pressure (DBP), and pulse pressure (PP) constitutes a risk factor for EPVSs within the basal ganglia and centrum semiovale^23^. Another study focusing on participants with cardiovascular risk factors reported a positive correlation between 24-h ambulatory blood pressure variability and the severity of basal ganglia EPVSs ^24^. Notably, previous research has focused primarily on the association of blood pressure levels and their variability with EPVSs, without addressing the impact of the duration of blood pressure control. To our knowledge, this is one of the first studies to investigate the association between SBP-TTR, a composite metric integrating the duration of blood pressure control, blood pressure level, and long-term blood pressure variability. More than ten years of follow-up data from the Kailuan community cohort revealed that a high SBP-TTR reduces the risk of EPVSs. The underlying mechanism may involve long-term stable blood pressure control to maintain cerebrospinal fluid circulation homeostasis and ensure proper metabolic clearance function of the glymphatic system, thereby promoting normal fluid flow within the PVSs. These findings suggest that early intervention targeting the SBP-TTR may effectively protect glymphatic system function.

This study confirmed that a high SBP-TTR is associated with a reduced risk of WMHs in the hypertensive population, independent of SBP variability. WMHs are an important neuroimaging marker of CSVD, reflecting the integrity of the brain parenchyma^25^. Their pathological progression is closely linked to neurological dysfunctions such as cognitive impairment, dementia, and stroke^26^. The pathological mechanism of WMHs may involve chronic hypoperfusion secondary to microvascular damage, leading to demyelination and microstructural changes in the white matter, ultimately resulting in radiologically visible WMHs^27^. In a community-based population, Li et al. reported that elevated SBP was significantly associated with increased total WMH volume^28^. A study by Middelaar et al. involving 122 elderly hypertensive patients (aged 70–78 years) indicated that increased SBPV was independently associated with the degree of WMH progression (OR=0.03, 95% CI: 0.00–0.05) ^29^(. A multicenter randomized clinical trial involving hypertensive patients aged ≥50 years revealed that the intensive blood pressure control group (SBP <120 mmHg) had a significantly smaller increase in WMH volume than the standard control group (SBP <140 mmHg) did (0.92 cm³ vs. 1.45 cm³)^30^. Notably, previous studies have not investigated the impact of the duration of blood pressure control on WMHs, and some studies have been limited by the age of the cohort (elderly population) or small sample sizes. On the basis of the Kailuan community cohort, which includes adults aged 30.5–81.8 years, this study confirmed that a high SBP-TTR is independently associated with a reduced risk of WMHs. The pathological WMH mechanism may involve chronic hypoperfusion induced by microvascular damage. High-quality, long-term blood pressure control may mitigate vascular endothelial injury and improve cerebral blood perfusion, thereby delaying WMH progression.

## 4. Limitations

Although this study employed rigorous methodological approaches in terms of both design and analytical procedures, several limitations should be acknowledged. First, the cross-sectional design: The assessment of CSVD neuroimaging markers relied exclusively on cranial MRI data obtained during the seventh follow-up survey. Consequently, this precludes causal inference regarding the association between SBP-TTR and CSVD incidence or progression, demonstrating only an association. Second, the cohort was recruited solely from the Kailuan community-based population in Tangshan, China. The participants’ lifestyle factors, genetic predispositions, and healthcare access exhibit regional specificity, potentially limiting the generalizability of the findings to broader ethnic or geographically diverse populations.

## 5. Conclusion

This study demonstrated that in hypertensive patients, the SBP-TTR is inversely associated with both EPVSs and WMHs. High-quality blood pressure management may serve as an effective strategy for controlling the progression of CSVD. SBP-TTR may be considered an objective indicator for evaluating the quality of blood pressure control in community-based hypertension management programs.

## Abbreviations

SBP-TTR: Systolic Blood Pressure Time in Target Range
CSVD: Cerebral Small Vessel Disease
LAs: Lacunes
CMBs: Cerebral Microbleeds
EPVSs: Enlarged Perivascular Spaces
WMHs: White Matter Hyperintensities
BMI: Body Mass Index
FBG: Fasting Blood Glucose
LDL-C: Low-Density Lipoprotein Cholesterol
HDL-C: High-Density Lipoprotein Cholesterol
TGs: Triglycerides
TC: Total Cholesterol
MRI: Magnetic Resonance Imaging
T1WI: T1-Weighted Imaging
T2WI: T2-Weighted Imaging
FLAIR: Fluid Attenuated Inversion Recovery
SWI: Susceptibility-Weighted Imaging
DWI: Diffusion-Weighted Imaging
SD: Standard Deviation
CV: Coefficient of Variation
OR: Odds Ratio
CI: Confidence Interval
ANOVA: Analysis of Variance
BPV: Blood Pressure Variability
SBPV: Systolic Blood Pressure Variability
SBP-STD: Standard Deviation of Systolic Blood Pressure
SBP-CV: Coefficient of Variation of Systolic Blood Pressure
DBP: Diastolic Blood Pressure
PP: Pulse Pressure
PVWMHs: Periventricular White Matter Hyperintensities
DWMHs: Deep White Matter Hyperintensities
PVSs: Perivascular Spaces

## Data Availability

Data can be obtained with the author’s consent.

## Acknowledgements

We sincerely express our gratitude to all participants in the Kailuan Study, as well as members of Kailuan General Hospital and its affiliated hospitals.

## Supplementary material

Supplementary material is available at Hypertension online

## References

1. Markus HS, de Leeuw FE. Cerebral small vessel disease: Recent advances and future directions. International journal of stroke: official journal of the International Stroke Society. Jan 2023;18(1):4–14. doi:10.1177/17474930221144911

2. Charidimou A, Pantoni L, Love S. The concept of sporadic cerebral small vessel disease: A road map on key definitions and current concepts. International journal of stroke: official journal of the International Stroke Society. Jan 2016;11(1):6–18. doi:10.1177/1747493015607485

3. Wardlaw JM, Smith C, Dichgans M. Small vessel disease: mechanisms and clinical implications. The Lancet Neurology. Jul 2019;18(7):684–696. doi:10.1016/s1474-4422(19)30079-1

4. Jacob MA, Cai M, van de Donk V, et al. Cerebral Small Vessel Disease Progression and the Risk of Dementia: A 14-Year Follow-Up Study. The American journal of psychiatry. Jul 1 2023;180(7):508–518. doi:10.1176/appi.ajp.20220380

5. Mills KT, Stefanescu A, He J. The global epidemiology of hypertension. Nature reviews Nephrology. Apr 2020;16(4):223–237. doi:10.1038/s41581-019-0244-2

6. Klarenbeek P, van Oostenbrugge RJ, Rouhl RP, Knottnerus IL, Staals J. Ambulatory blood pressure in patients with lacunar stroke: association with total MRI burden of cerebral small vessel disease. Stroke. Nov 2013;44(11):2995–9. doi:10.1161/strokeaha.113.002545

7. Parati G, Bilo G, Kollias A, et al. Blood pressure variability: methodological aspects, clinical relevance and practical indications for management - a European Society of Hypertension position paper ∗. Journal of hypertension. Apr 1 2023;41(4):527–544. doi:10.1097/hjh.0000000000003363

8. Zhao X, Hui Y, Li J, et al. Higher Long-Term Visit-to-Visit Blood Pressure Variability Is Associated With Severe Cerebral Small Vessel Disease in the General Population. *Journal of clinical hypertension (Greenwich*, Conn). Jan 2025;27(1):e14943. doi:10.1111/jch.14943

9. Ma Y, Song A, Viswanathan A, et al. Blood Pressure Variability and Cerebral Small Vessel Disease: A Systematic Review and Meta-Analysis of Population-Based Cohorts. Stroke. Jan 2020;51(1):82–89. doi:10.1161/strokeaha.119.026739

10. Fatani N, Dixon DL, Van Tassell BW, Fanikos J, Buckley LF. Systolic Blood Pressure Time in Target Range and Cardiovascular Outcomes in Patients With Hypertension. Journal of the American College of Cardiology. Mar 16 2021;77(10):1290–1299. doi:10.1016/j.jacc.2021.01.014

11. Doumas M, Tsioufis C, Fletcher R, Amdur R, Faselis C, Papademetriou V. Time in Therapeutic Range, as a Determinant of All-Cause Mortality in Patients With Hypertension. Journal of the American Heart Association. Nov 3 2017;6(11)doi:10.1161/jaha.117.007131

12. Schmitt L, Speckman J, Ansell J. Quality assessment of anticoagulation dose management: comparative evaluation of measures of time-in-therapeutic range. Journal of thrombosis and thrombolysis. Jun 2003;15(3):213–6. doi:10.1023/b:Thro.0000011377.78585.63

13. Chung SC, Pujades-Rodriguez M, Duyx B, et al. Time spent at blood pressure target and the risk of death and cardiovascular diseases. PloS one. 2018;13(9):e0202359. doi:10.1371/journal.pone.0202359

14. Han X, Liu S, Zhou X, Chen S, Wu S, Yang Q. Systolic Blood Pressure Time in Target Range and Cardiovascular Disease and Premature Death. JACC Asia. Dec 2024;4(12):987–996. doi:10.1016/j.jacasi.2024.09.002

15. Li S, Jiang C, Wang Y, et al. Systolic Blood Pressure Time in Target Range and Cognitive Outcomes: Insights From the SPRINT MIND Trial. Hypertension (Dallas, Tex: 1979). Aug 2023;80(8):1628–1636. doi:10.1161/hypertensionaha.122.20711

16. Rudilosso S, San Román L, Blasco J, Hernández-Pérez M, Urra X, Chamorro Á. Evaluation of white matter hypodensities on computed tomography in stroke patients using the Fazekas score. Clinical imaging. Nov-Dec 2017;46:24–27. doi:10.1016/j.clinimag.2017.06.011

17. Staals J, Makin SD, Doubal FN, Dennis MS, Wardlaw JM. Stroke subtype, vascular risk factors, and total MRI brain small-vessel disease burden. Neurology. Sep 30 2014;83(14):1228–34. doi:10.1212/wnl.0000000000000837

18. Jiang L, Cai X, Yao D, et al. Association of inflammatory markers with cerebral small vessel disease in community-based population. Journal of neuroinflammation. May 6 2022;19(1):106. doi:10.1186/s12974-022-02468-0

19. Javierre-Petit C, Schneider JA, Kapasi A, et al. Neuropathologic and Cognitive Correlates of Enlarged Perivascular Spaces in a Community-Based Cohort of Older Adults. Stroke. Sep 2020;51(9):2825–2833. doi:10.1161/strokeaha.120.029388

20. Taoka T, Naganawa S. Neurofluid Dynamics and the Glymphatic System: A Neuroimaging Perspective. Korean journal of radiology. Nov 2020;21(11):1199–1209. doi:10.3348/kjr.2020.0042

21. Raicevic N, Forer JM, Ladrón-de-Guevara A, et al. Sizes and shapes of perivascular spaces surrounding murine pial arteries. Fluids and barriers of the CNS. Jul 17 2023;20(1):56. doi:10.1186/s12987-023-00454-z

22. Berezuk C, Ramirez J, Gao F, et al. Virchow-Robin Spaces: Correlations with Polysomnography-Derived Sleep Parameters. Sleep. Jun 1 2015;38(6):853–8. doi:10.5665/sleep.4726

23. Shi H, Cui L, Hui Y, et al. Enlarged Perivascular Spaces in Relation to Cumulative Blood Pressure Exposure and Cognitive Impairment. Hypertension (Dallas, Tex: 1979). Oct 2023;80(10):2088–2098. doi:10.1161/hypertensionaha.123.21453

24. Yang S, Qin W, Yang L, et al. The relationship between ambulatory blood pressure variability and enlarged perivascular spaces: a cross-sectional study. BMJ open. Aug 21 2017;7(8):e015719. doi:10.1136/bmjopen-2016-015719

25. Bonkhoff AK, Hong S, Bretzner M, et al. Association of Stroke Lesion Pattern and White Matter Hyperintensity Burden With Stroke Severity and Outcome. Neurology. Sep 27 2022;99(13):e1364–e1379. doi:10.1212/wnl.0000000000200926

26. Guo W, Shi J. White matter hyperintensities volume and cognition: A meta-analysis. Frontiers in aging neuroscience. 2022;14:949763. doi:10.3389/fnagi.2022.949763

27. Bahrani AA, Powell DK, Yu G, Johnson ES, Jicha GA, Smith CD. White Matter Hyperintensity Associations with Cerebral Blood Flow in Elderly Subjects Stratified by Cerebrovascular Risk. Journal of stroke and cerebrovascular diseases: the official journal of National Stroke Association. Apr 2017;26(4):779–786. doi:10.1016/j.jstrokecerebrovasdis.2016.10.017

28. Li X, Hui Y, Shi H, et al. Association of blood pressure with brain perfusion and structure: A population-based prospective study. European journal of radiology. Aug 2023;165:110889. doi:10.1016/j.ejrad.2023.110889

29. van Middelaar T, Richard E, Moll van Charante EP, van Gool WA, van Dalen JW. Visit-to-Visit Blood Pressure Variability and Progression of White Matter Hyperintensities Among Older People With Hypertension. Journal of the American Medical Directors Association. Sep 2019;20(9):1175–1177.e1. doi:10.1016/j.jamda.2019.04.003

30. Nasrallah IM, Pajewski NM, Auchus AP, et al. Association of Intensive vs Standard Blood Pressure Control With Cerebral White Matter Lesions. Jama. Aug 13 2019;322(6):524–534. doi:10.1001/jama.2019.10551

